# Immunogenicity and safety of LP.8.1 variant-containing mRNA COVID-19 vaccines

**DOI:** 10.64898/2026.02.24.26346954

**Authors:** Amparo L. Figueroa, Kimball Johnson, Robert Springer, Jarrett Lowe, Ann Cripple, Darin K Edwards, Wenqin Xu, Xin Cao, Veronica Urdaneta, Bethany Girard, Arshan Nasir, David C. Montefiori, Spyros Chalkias

**Author notes:** Corresponding author: Spyros Chalkias, Moderna, Inc., 325 Binney Street, Cambridge, MA, USA 02142.

## Abstract

**Background:** The SARS-CoV-2 LP.8.1 subvariant was incorporated into the 2025-2026 U.S. COVID-19 vaccines (mRNA-1273.251 and mRNA-1283.251). We evaluated immunogenicity and safety of these vaccines against vaccine-matched and emerging variants in individuals aged ≥65 and those aged 12-64 years at high-risk of severe COVID-19.

**Methods:** Data were generated from: (1) two independent, ongoing, phase 3b/4, open-label, single-arm studies in which participants received a single dose of 50-µg mRNA-1273.251 (n=103; median age, 64.0 years) or 10-µg mRNA-1283.251 (n=172, median age, 59.0 years) and followed through Day 29 post-vaccination; neutralizing antibodies (nAb) were measured at baseline (Day 1) and Day 29 using a pseudovirus neutralization assay against the vaccine-matched LP.8.1 variant; (2) Day 29 immunogenicity against circulating variants (BA.3.2.2, XFG, and NB.1.8.1) was assessed in a randomly selected subset; and (3) immune-escape potential was estimated using predictive modeling. Unsolicited adverse events (AEs), including serious AEs, leading to study withdrawal, and those of special interest, were monitored.

**Results:** Both vaccines elicited robust nAbs at Day 29 against LP.8.1 (geometric mean fold-rise from baseline: 12-64 years, mRNA-1273.251, 26.3; mRNA-1283.251, 53.0; ≥65 years, mRNA-1273.251, 15.4; mRNA-1283.251, 36.7) and circulating variants. Model-based estimates with mRNA-1273.251 were consistent with clinical data and indicated the highest responses against LP.8.1 and lower responses against BA.3.2.2. No vaccine-related AEs were reported in either study.

**Conclusions:** mRNA-1273.251 and mRNA-1283.251 were well tolerated through Day 29 and elicited robust nAbs against vaccine-matched and circulating variants. In predictive models, BA.3.2.2 had the highest relative risk of immune escape following mRNA-1273.251 vaccination.

**SUMMARY:** LP.8.1-containing mRNA-1273.251 and mRNA-1283.251 vaccines given as a single dose were well tolerated and induced robust Day 29 neutralizing antibodies against LP.8.1 and circulating variants.

## INTRODUCTION

The SARS-CoV-2 JN.1 lineage, first detected in late 2023, evolved and diversified throughout late 2024 and 2025, driving successive waves of subvariants with potential for enhanced transmissibility and immune escape [1, 2]. One such subvariant, LP.8.1, carries nine additional spike (S) mutations relative to JN.1 [3]. LP.8.1 rose to dominance in several regions, including the United States, in late 2024 and early 2025, accounting for 61% of sequenced variants in May 2025 before gradually declining as newly emerging JN.1 subvariants increased in frequency [4]. Based on global genomic surveillance during 2024-2025, LP.8.1 was selected for inclusion in the 2025–2026 U.S. COVID-19 vaccine composition [5]. As LP.8.1 waned, recombination between two JN.1-derived subvariants gave rise to XFG, which is currently the dominant circulating variant in the United States [6, 7]. In parallel, NB.1.8.1, another variant with a JN.1-like S protein has been growing steadily, predominantly in Asian countries [6]. More recently, BA.3.2, a new long-branch variant, has been increasing in parts of Europe and Australia, with travel-related cases detected elsewhere [6]. BA.3.2, descended from the ancestral BA.3 variant that circulated briefly in early 2022, is a genetically divergent strain that likely evolved via prolonged evolution in immunocompromised hosts during this 3 year period [8].

Timely characterization of immune responses elicited by updated vaccines remains important as new variants emerge and antigenically diversify. To support this goal, two phase 3b/4 trials (NCT06585241 and NCT07089706) were initiated to generate clinical data for updated variant-containing formulations of mRNA-1273 (SPIKEVAX, Moderna, Inc.) and mRNA-1283 (mNEXSPIKE, Moderna, Inc.).

mRNA-1273 COVID-19 vaccine, developed using the Sponsor’s mRNA-lipid nanoparticle (LNP) based platform, contains a single mRNA encoding the full-length SARS-CoV-2 S protein, which enables host-cell viral access [9, 10]. mRNA-1283 is a COVID-19 vaccine that encodes only the SARS-CoV-2 S protein regions most critical for eliciting protective neutralizing antibodies (nAbs), the receptor-binding domain (RBD) and N-terminal domain (NTD), which not only strengthen elicited immune responses, but also allow for mRNA-1283 to be administered at lower doses and confer greater stability, improving storage and shelf-life [11–13]. mRNA-1273 and mRNA-1283 are approved in the United States for individuals aged ≥65 years and for those aged 6 months through 64 years (mRNA-1273) or 12 through 64 years (mRNA-1283) with at least one underlying condition that places them at high risk for severe COVID-19 outcomes [11, 12, 14].

Here, we report Day 29 immunogenicity and safety results from two independent phase 3b/4 studies evaluating the current (JN.1-lineage) 2025–2026 recommended variant-containing formulations of the mRNA-1273.251 and mRNA-1283.251 vaccines in individuals aged ≥65 years and in individuals aged ≥12 to <65 years with risk factors for severe COVID-19. In addition, we also describe the predicted immune escape of emerging variants based on our predictive modeling.

## METHODS

Data described herein were generated via three analyses: (1) two independent, ongoing, phase 3b/4, open-label, single-arm clinical studies evaluating immunogenicity and safety of LP.8.1-containing mRNA vaccines; (2) fit-for-purpose assays and exploratory analyses of Day 29 immunogenicity against emerging variants (BA.3.2.2, XFG, and NB.1.8.1), conducted in a randomly-selected subset of participants from the phase 3b/4 trials; and (3) *in silico* prediction of immune escape using a previously described risk-calculator [15].

### Phase 3b/4 Trials

#### Study design and participants

Two independent, ongoing, phase 3b/4, open-label, single-arm studies evaluated the immunogenicity and safety of the 2025-2026 LP.8.1-containing formulations of mRNA-1273 and mRNA-1283 vaccines (mRNA-1273.251 and mRNA-1283.251, respectively) given as a single dose. Each study was conducted at five sites in the United States (mRNA-1273.251: NCT06585241; mRNA-1283.251: NCT07089706). Immunogenicity assessments were evaluated on Day 1 (baseline) and Day 29; safety was assessed through Day 29.

Eligible participants were adults aged ≥65 years or individuals aged ≥12 to <65 years with at least one underlying condition associated with increased risk for severe COVID-19 outcomes and who provided written informed consent. Full inclusion/exclusion criteria are provided in the **Supplement**. Participants received a single dose of either mRNA-1273.251 (50 µg) or mRNA-1283.251 (10 µg) on Day 1; the final study visit occurred on Day 29.

Both studies were conducted in accordance with the protocol and consensus and ethical principles derived from international guidelines, including the Declaration of Helsinki, Council for International Organizations of Medical Sciences, applicable International Council for Harmonization Good Clinical Practice guidelines, and relevant laws and regulations. The protocol was approved by the Advarra Institutional Review Board before study initiation, and written informed consent was received by all participants before enrollment. Participants and the public were not involved in the design, conduct, or reporting of this trial.

### Study vaccines

Both mRNA-1273.251 and mRNA-1283.251 are LNP-encapsulated mRNA vaccines. mRNA-1273.251 encodes the full-length prefusion-stabilized LP.8.1 variant S glycoprotein, and mRNA-1283.251 encodes the RBD and NTD segments of the SARS-CoV-2 S protein of the LP.8.1 variant. Participants received a single 0.5-mL dose of mRNA-1273.251 (50 µg) or 0.20 mL dose of mRNA-1283.251 (10 µg) intramuscularly (deltoid) on Day 1.

### Objectives

The primary objective was to evaluate the nAb response elicited by mRNA-1273.251 or mRNA-1283.251 against the vaccine-matched LP.8.1 variant at Day 29. The secondary objective was to evaluate the safety of each vaccine through Day 29.

### Immunogenicity assessments

Blood samples for immunogenicity assessments were collected on Days 1 (baseline) and 29 (end of study). Primary immunogenicity endpoints included the geometric mean titer (GMT) at Days 1 and 29, geometric mean fold rise (GMFR) at Day 29 over Day 1, and seroresponse rate (SRR) of serum nAb against the vaccine-matched SARS-CoV-2 variant measured by pseudovirus-based neutralization assays (detailed in the **Supplement**).

### Safety assessments

Safety endpoints included unsolicited adverse events (AEs), serious AEs (SAEs), AEs leading to study withdrawal, and AEs of special interest (AESIs) from Day 1 through Day 29.

### Fit-For-Purpose Assays and Exploratory Analyses

A fit-for-purpose approach was applied to enable rapid assay development and sample testing for newly emerging variants. An ad hoc population comprising a randomly selected subset of 56 participants from the two phase 3b/4 trials was used for variant neutralization testing against emerging variants (BA.3.2.2, XFG, and NB.1.8.1). GMT at Days 1 and 29 and GMFR at Day 29 over Day 1 of serum nAbs against circulating SARS-CoV-2 variants of concern or new variants (LP.8.1, BA.3.2.2, XFG, NB.1.8.1) were measured by pseudovirus-based neutralization assays (detailed in the **Supplement**).

### Predictive Model of Immune Escape

To estimate immune escape potential of emerging SARS-CoV-2 variants, we applied an *in silico* risk calculator previously described by Nasir, et al. [15]. This risk calculator uses statistical modeling, trained on a large panel of pseudotyped SARS-CoV-2 variants assayed for neutralization activity against clinical sera, and integrates S antigenic and antibody escape profiles from deep mutational scanning studies [16–19]. The model generates a quantitative prediction of variant immune escape based on newly observed RBD mutations relative to prior vaccine strains. For each variant, the calculator produces an estimate of the relative fold reduction in ID50 neutralization titers compared to the corresponding vaccine strain, representing the expected decrease in serum neutralization potency.

### Statistical Analyses

#### Phase 3b/4 trials

No hypothesis testing was performed, and all analyses were descriptive. A total of 104 participants received the mRNA-1273.251 and 172 participants received the mRNA-1283.251. Primary immunogenicity assessments were based on the per-protocol immunogenicity set (PPIS), which included all participants that received the study intervention, had negative reverse transcription polymerase chain reaction (RT-PCR) tests on Days 1 and 29, and had no major protocol deviations that impacted key data.

The 95% confidence intervals (CIs) for GMT and GMFR were calculated based on the *t*-distribution of the log transformed values and then back transformed to the original scale for presentation. The SRR, which is the percentage of participants with seroresponse on Day 29, was provided along with the 2-sided 95% CI estimated using the Clopper-Pearson method. Antibody values below the lower limit of quantification (LLOQ) were replaced by 0.5xLLOQ. Values greater than the upper limit of quantification (ULOQ) were replaced by the ULOQ.

All safety analyses were based on the safety set (all enrolled participants who received study vaccination).

### Fit-for-purpose exploratory analyses

The ad hoc population comprised a subset of participants stratified by age group (individuals aged ≥65 years, and those aged ≥12 to <65 years with at least one underlying condition that put them at high risk for severe COVID-19 outcomes). GMTs at Days 1 and 29, and GMFR at Day 29 over Day 1 of serum nAbs against circulating SARS-CoV-2 variants of concern or new variants (LP.8.1, BA.3.2.2, XFG, NB.1.8.1) were calculated. Limit of detection (LOD) was used for analyses across all four variants. This contrasts with the LP.8.1 assay, which was fully validated prior to use and employed LLOQ and ULOQ derived through a formal validation process. Antibody values below the LOD were imputed as 0.5×LOD. Values above the ULOQ (applied only to the LP.8.1 assay) were not capped.

## RESULTS

### Phase 3b/4 Trials

#### Participants

At the data cutoff date (mRNA-1273.251: October 23, 2025; mRNA-1283.251: October 31, 2025), 104 participants were independently enrolled in the mRNA-1273.251 study (103 received mRNA-1273.251; median age, 64.0 years; 52.4% aged ≥12 to <65 years; 53.4% female; 40.8% white), and 172 participants were enrolled in the mRNA-1283.251 study and received the study vaccination (median age, 59.0 years; 64.0% aged ≥12 to <65 years; 61.6% female; 53.5% white) (**Figure 1**; **Table 1**).

**Figure 1.**
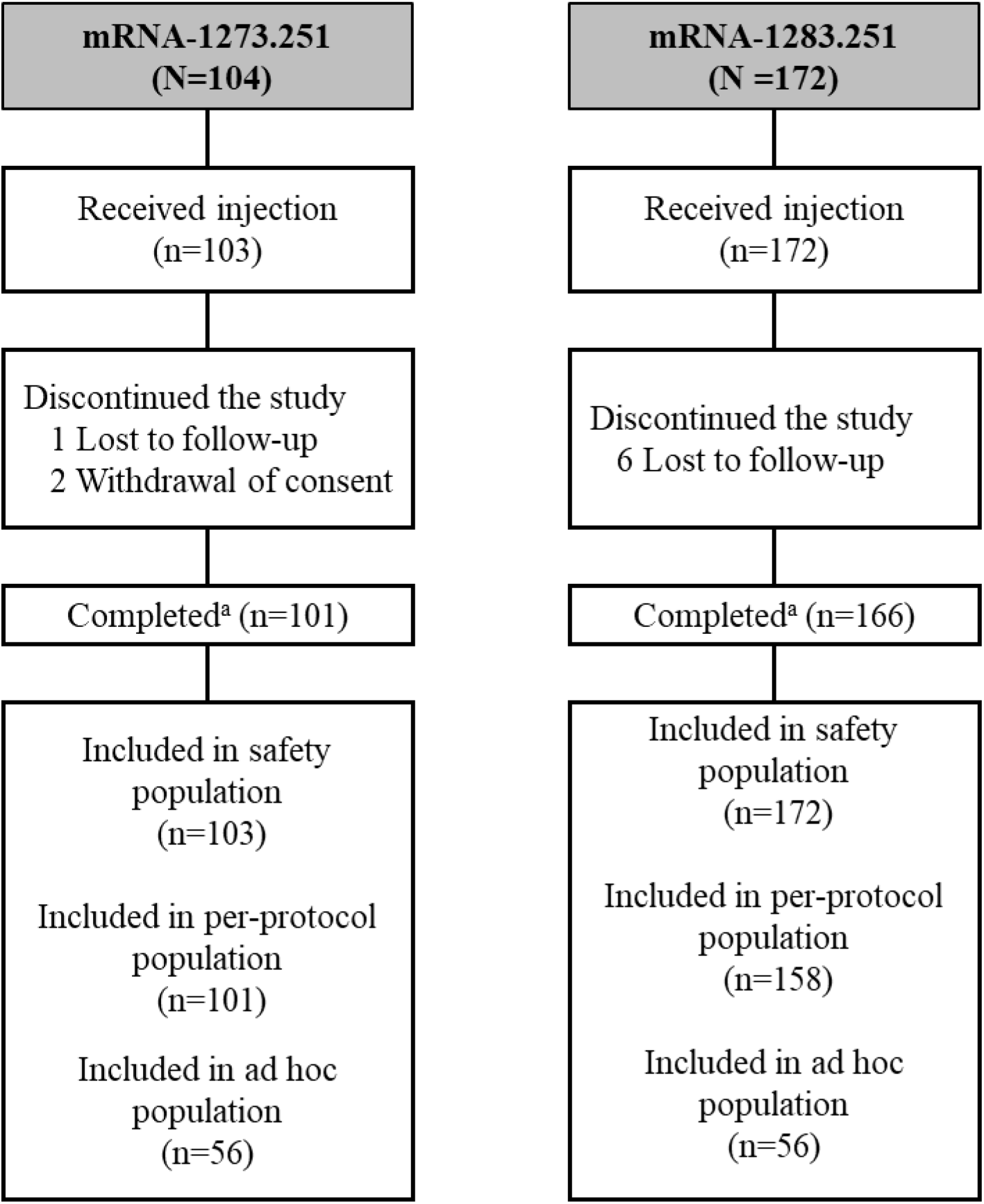
Participant disposition. RT-PCR; reverse transcription polymerase chain reaction. All participants who were enrolled and received the study vaccination were included in the Safety Set. The immunogenicity Per-Protocol population included all enrolled participants who received the study intervention, had negative RT-PCR tests on Days 1 and 29, and had no major protocol deviations that impacted key data. ^a^Participants were considered to have completed the study if they had completed the last scheduled procedure on Day 29.

**Table 1.**
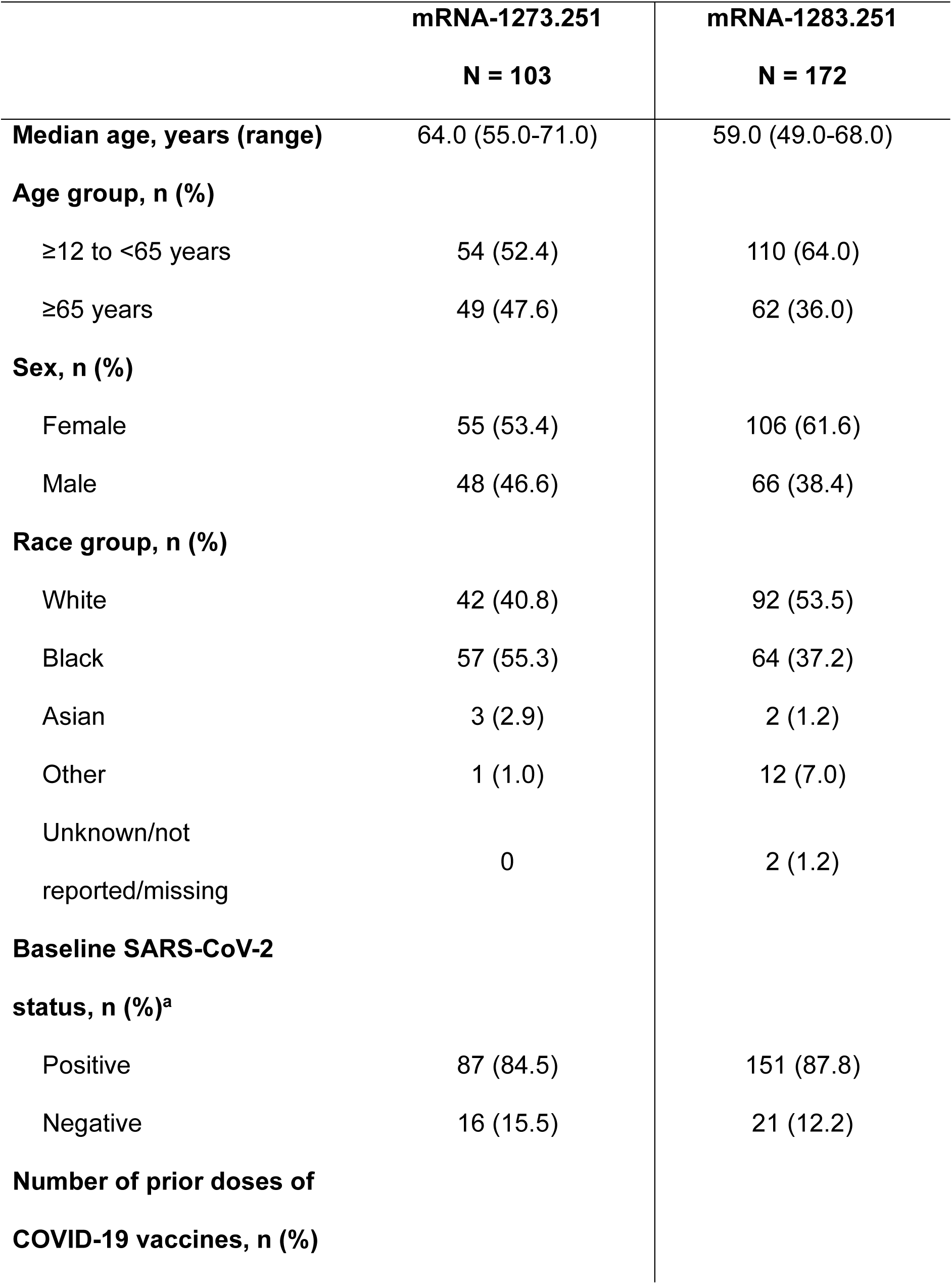
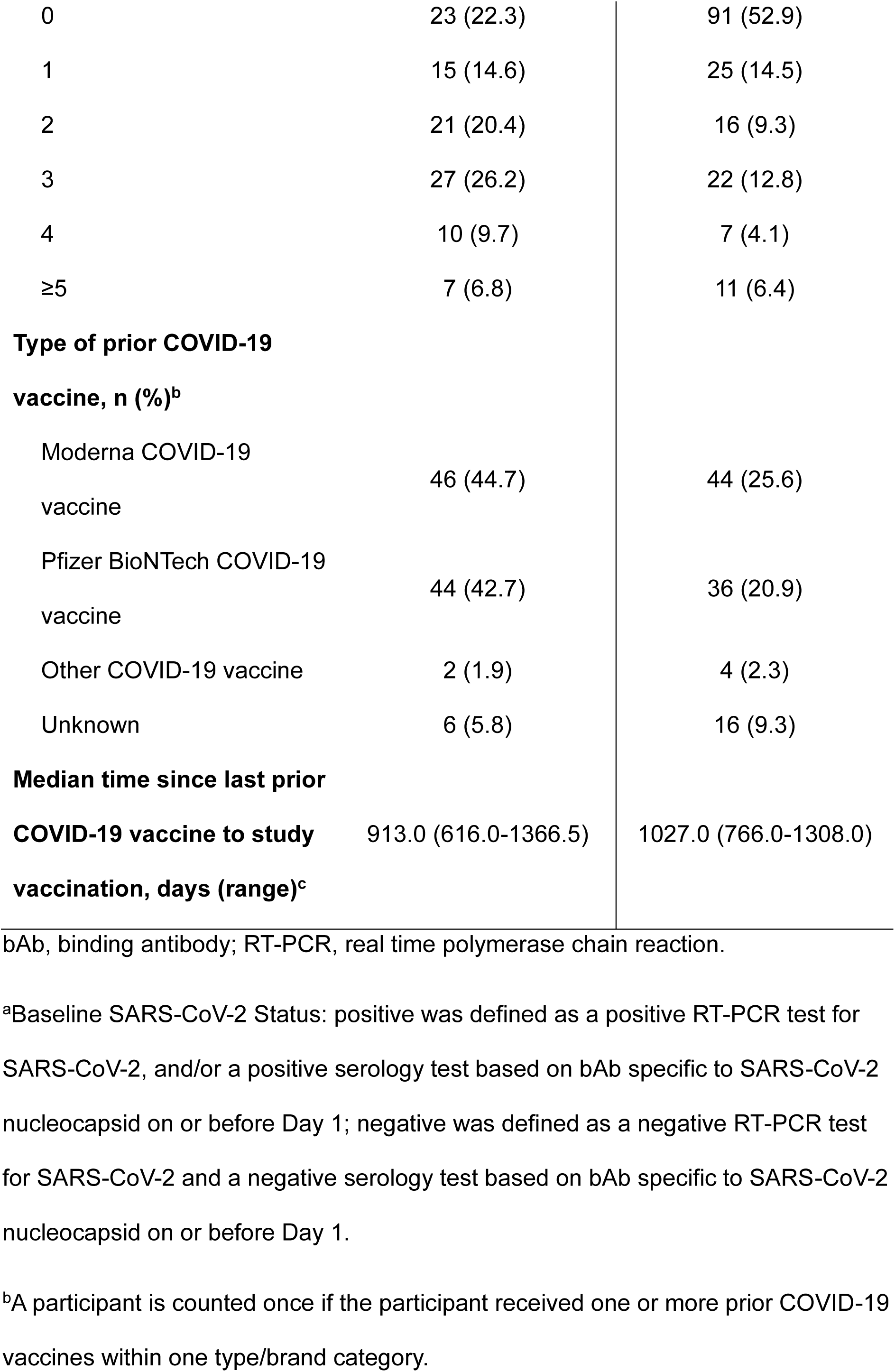
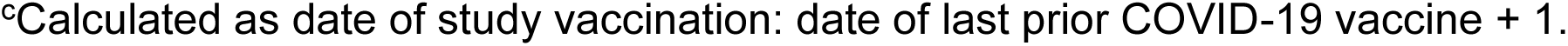
Baseline demographic and clinical characteristics.

Baseline SARS-CoV-2 status was determined by virologic (RT-PCR) and/or serologic (anti-nucleocapsid binding antibody) evidence of SARS-CoV-2 infection on or before Day 1. In the mRNA-1273.251 study, 87 (84.5%) participants had a positive baseline SARS-CoV-2 status; PPIS included 101 (98.1%) participants (2 participants were excluded due to a missing or positive SARS-CoV-2 RT-PCR result on Day 29). In the mRNA-1283.251 study, 151 (87.8%) participants had a positive baseline SARS-CoV-2 status; PPIS included 158 (91.9%) participants (14 participants were excluded due to a positive SARS-CoV-2 RT-PCR result on Day 29).

Overall, in the mRNA-1273.251 study, 14.6%-26.2% of participants received one to three prior doses of COVID-19 vaccines. The median time since last prior COVID-19 vaccine to study vaccination was 913.0 days (range, 616.0-1366.5). In the mRNA-1283.251 study, 12.8%-14.5% of participants received 1 or 3 prior doses COVID-19 vaccines. The median time since last prior COVID-19 vaccine to study vaccination was 1027.0 days (range, 766.0-1308.0).

### Immunogenicity against the vaccine matched variant

Both vaccines elicited robust increases in nAb responses at Day 29 relative to baseline against the matched variant (LP.8.1) in participants aged ≥12 to <65 years (GMFR, mRNA-1273.251: 26.3 [95% CI, 15.6-44.3]; mRNA-1283.251: 53.0 [95% CI, 36.7-76.4]), and in participants aged ≥65 years (GMFR, mRNA-1273.251: 15.4 [95% CI, 9.5-25.0]; mRNA-1283.251: 36.7 [95% CI, 23.1-58.4]; **Figure 2, Table S1**).

**Figure 2.**
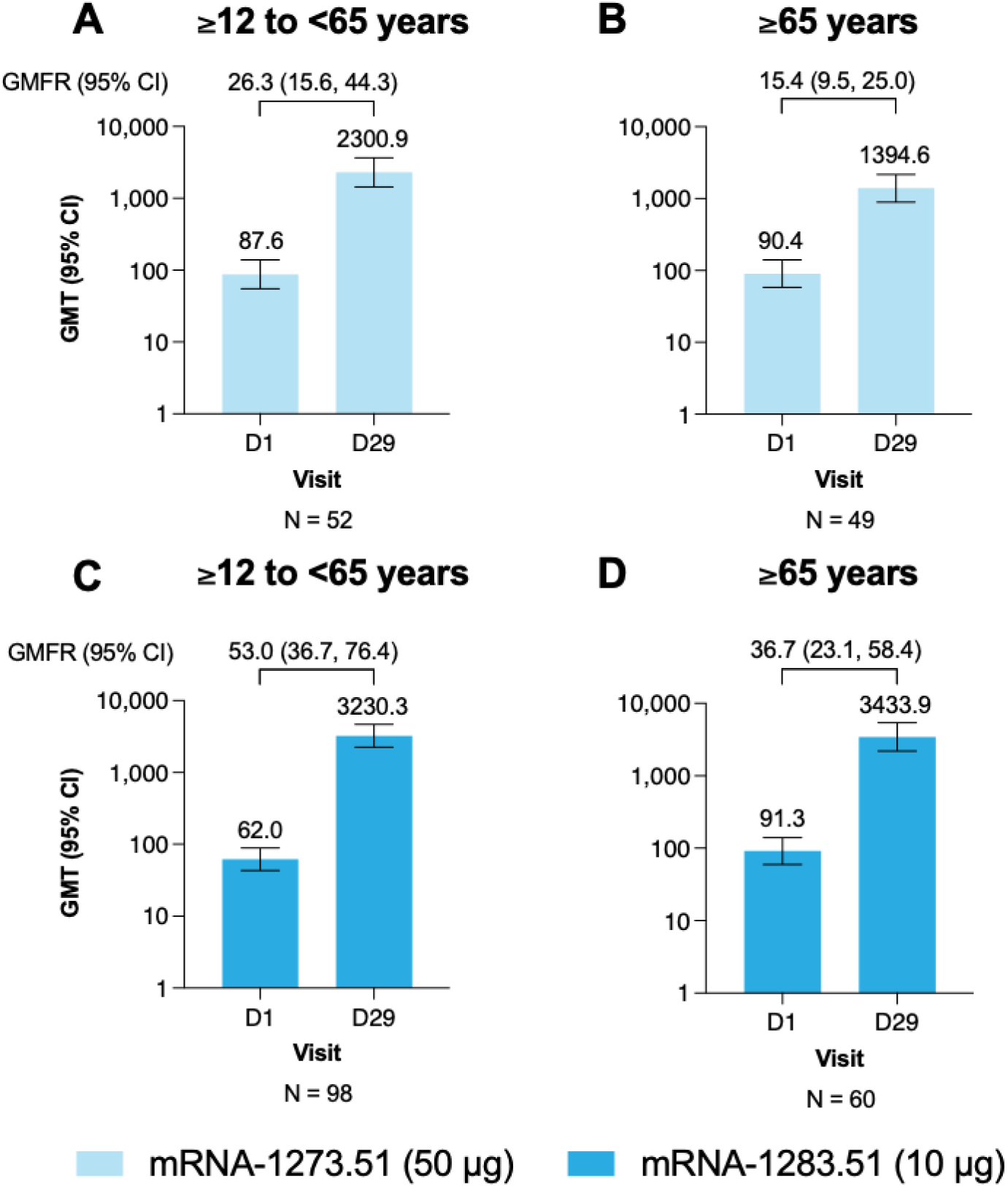
Neutralizing antibody titers against LP.8.1 by age group at baseline and Day 29, per protocol primary analysis. GMT, geometric mean titer; LLOQ, lower limit of quantification; nAb, neutralizing antibody; ULOQ, upper limit of quantification. Responses elicited by mRNA-1273.251 in (A) participants aged ≥12 to <65 years and (B) ≥65 years; and responses elicited by mRNA-1283.251 in (C) participants aged ≥12 to <65 years; and (D) ≥65 years. Data cutoff date for immunogenicity: October 23, 2025 (mRNA-1273.251); October 31, 2025 (mRNA-1283.251). Antibody values reported as <LLOQ are replaced by 0.5 x LLOQ. Values >ULOQ are replaced by the ULOQ. mRNA-1273.251 nAb LLOQ 35, ULOQ 38606. mRNA-1283.251 nAb LLOQ 35, ULOQ 38606. 95% CI is calculated based on the t-distribution of the log-transformed values for GMT level, then back transformed to the original scale for presentation.

### Safety

At 28 days post-vaccination, no SAEs, AESIs, AEs that led to study discontinuation or deaths were reported for either vaccine. One AE (paraesthesia) was reported in the mRNA-1273.251 study, and two AEs (hypersensitivity, n = 1; tooth abscess, n = 1) were reported in the mRNA-1283.251 study; all AEs were considered unrelated to the study vaccine by the investigator.

### Fit-for-Purpose Exploratory Analyses

In a subset of randomly selected participants, both vaccines effectively cross-neutralized all tested emerging variants (BA.3.2.2, XFG, and NB.1.8.1) not encoded in the vaccines (**Figure 3**). At Day 29, after mRNA-1273.251 vaccination, GMTs were 1973.3 for LP.8.1, 823.6 for BA.3.2.2, 952.4 for XFG and 2064.7 for NB.1.8.1. The corresponding GMTs following mRA-1283.251 were 3827.2, 1549.3, 2050.6 and 3818.4, respectively.

**Figure 3.**
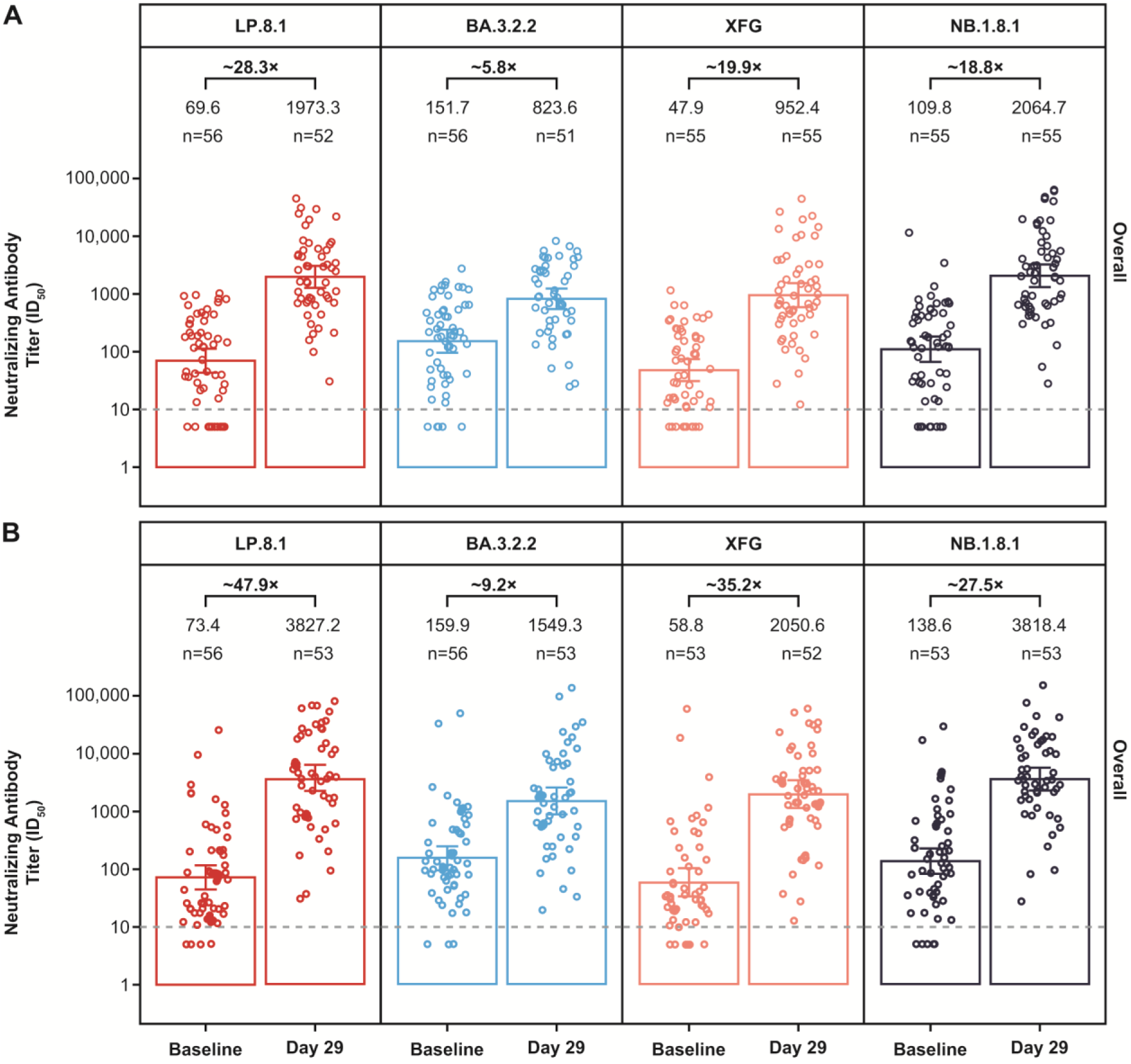
Neutralizing antibody responses elicited by (A) mRNA-1273.251 and (B) mRNA-1283.251 against vaccine-matched (LP.8.1) and emerging variants (BA.3.2.2, XFG, and NB.1.8.1), Fit-for-purpose analysis. nAb, neutralizing antibody; ID_50_, 50% inhibitory dose; LLOQ, lower limit of quantification; LOD, limit of detection; ULOQ, upper limit of quantification. Dashed lines represent LOD. A fit-for-purpose approach was applied to enable rapid assay development and sample testing for newly emerging variants. Accordingly, LOD was used for analyses across all four variants. This contrasts with the LP.8.1 assay, which was fully validated prior to use and employed LLOQ and ULOQ derived through a formal validation process. Antibody values below the LOD were imputed as 0.5×LOD. Values above the ULOQ were not capped; the ULOQ applied only to the LP.8.1 assay.

In participants aged ≥12 to <65 years and ≥65 years, post-vaccination nAb responses against the emerging variants BA.3.2.2, XFG, and NB.1.8.1 increased at Day 29 relative to baseline and were consistent across age groups (**Figure S1**). In both age groups, the highest and lowest titers for both vaccines were measured against NB.1.8.1 (GMT: mRNA-1273.251: 2182.8 [≥12 to <65 years] and 1956.9 [≥65 years]; mRNA-1283.251: 4090.7 [≥12 to <65 years] and 3573.3 [≥65 years]), and BA.3.2.2 (GMT: mRNA-1273.251: 825.9 [≥12 to <65 years] and 821.4 [≥65 years]; mRNA-1283.251: 1661.4 [≥12 to <65 years] and 1448.6 [≥65 years]), respectively. Compared with participants aged ≥65 years, younger participants receiving mRNA-1273.251 had slightly higher titers against all three emerging variants; younger participants receiving mRNA-1283.251 had higher titers against BA.3.2.2 and NB.1.8.1 compared with participants aged ≥65.

### Predictive Modeling of Immune Escape

The model-based matrix of predicted immune escape showed the greatest neutralization with mRNA-1273.251 LP.8.1 (7^th^ dose) against LP.8.1 variants (risk score of 1.0, indicating no reduction in ID_50_ nAb GMTs), with the lowest response predicted against the BA.3.2.2 variant (4.58-fold reduction in ID_50_ nAb GMTs), and modest neutralization against the XFG variant (2.62-fold reduction in ID_50_ nAb GMTs) (**Figure 4**).

**Figure 4.**
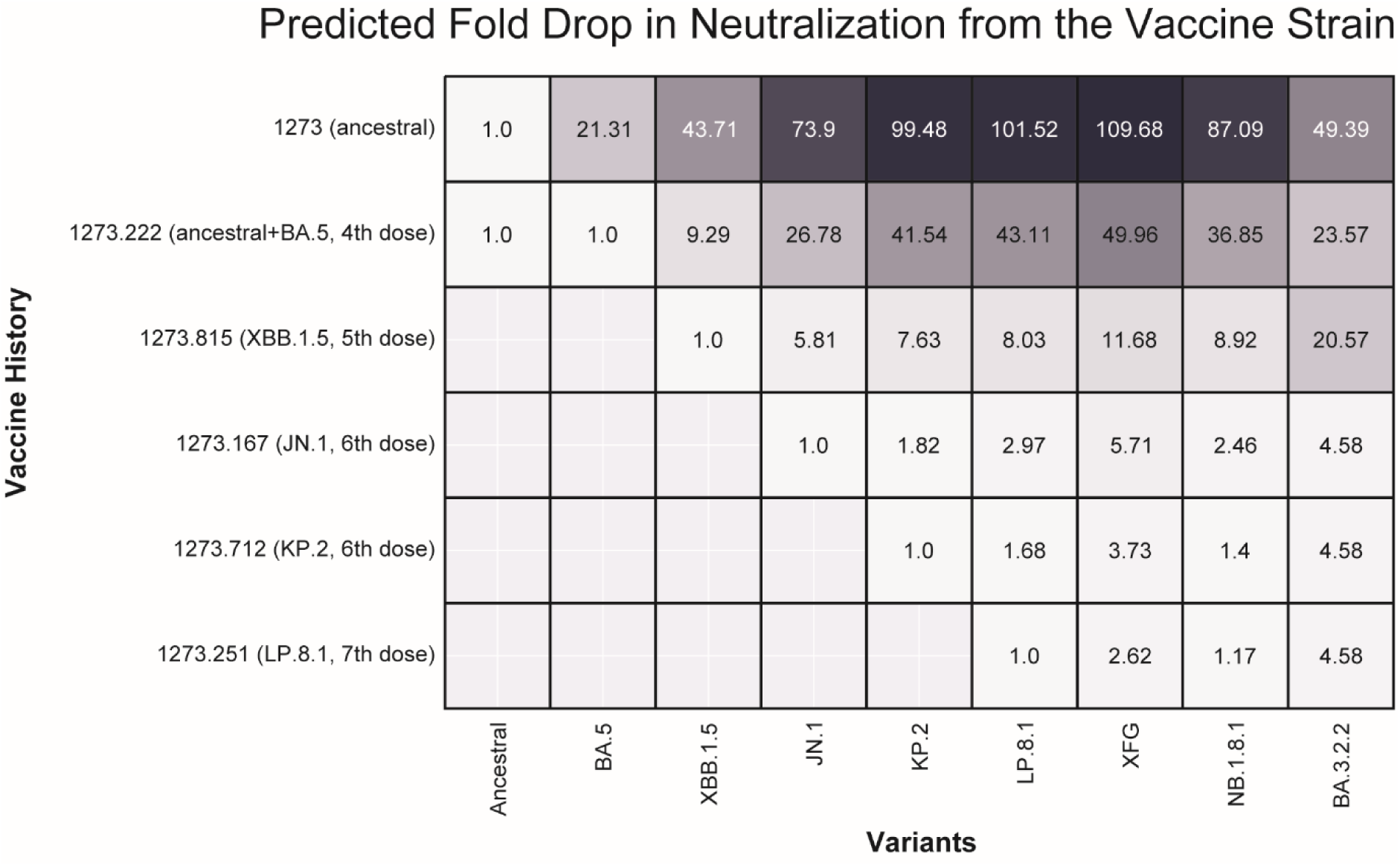
Predicted immune escape of SARS-CoV-2 variants across immunization backgrounds. nAb, neutralizing antibody; GMT, geometric mean titer; ID_50_, 50% inhibitory dose. Matrix showing model-estimated fold-reductions in ID_50_ nAb GMTs against the vaccine strain for each variant (x-axis) across different immunization backgrounds (y-axis). Each cell represents the predicted fold-reduction as estimated by the statistical model described in Nasir et al. [15]. Higher fold-reductions indicate greater predicted immune escape.

## DISCUSSION

In the two independent, ongoing phase 3b/4 studies in adults aged ≥65 years, and in individuals aged ≥12 to <65 years with risk factors for severe COVID-19, a single dose of mRNA.1273.251 and mRNA-1283.251 LP.8.1-containing vaccines was well tolerated and increased nAb responses against the vaccine-matched strain (LP.8.1). Both vaccines effectively neutralized all tested currently circulating variants at Day 29 across age groups. Moreover, the risk assessment model predicted the highest risk of immune escape for BA.3.2.2 (followed by XFG and NB.1.8.1). These estimates are consistent with the clinical data indicating potent immune responses following immunization with JN.1-containing mRNA vaccines against both closely related JN.1 strains (XFG and NB.1.8.1), and genetically distant BA.3.2.2 [20].

Taken together, these results suggest mRNA-1273.251 and mRNA-1283.251 LP.8.1-containing vaccines increased the immune responses against the vaccine strain and provided cross-variant neutralization against subvariants such as BA.3.2.2, XFG and NB.1.8.1. However, reduction in cross-neutralization may occur with newer variants, given the ongoing antigenic evolution and the potential immune-escape, as indicated by model-based estimates (**Figure 2**). The results also reinforce the importance of continued surveillance and periodic strain updates to reduce the impact of COVID-19, particularly in populations at increased risk for severe COVID-19 outcomes.

Limitations include lack of longer-term immunogenicity data for nAb responses. Although enrollment occurred at the same time, the results derive from two independent, open-label studies, and a single randomized study, which limits direct comparisons between vaccines. Each study also used distinct analytical populations: the PPIS comprised the predefined vaccine analysis population, whereas the fit-for-purpose analyses were designed to compare variants, not vaccines. While the primary and fit-for-purpose analyses used different thresholds for determining lower and upper limits of titers, the observed trends were consistent across both analyses and with prior evaluations.

In conclusion, LP.8.1-containing mRNA-1273.251 and mRNA-1283.251 vaccines increased nAb responses against the vaccine-matched variant and cross-neutralized other circulating variants.

## ACKNOWLEDGMENTS

Medical writing and editorial assistance were provided by Aliscia Daniels, PhD, and Shanel Dhani, PhD, of MEDiSTRAVA in accordance with Good Publication Practice (GPP 2022) guidelines, funded by Moderna, Inc., and under the direction of the authors.

## AUTHOR CONTRIBUTIONS

A.L.F., J.L., A.C., D.K.E., W.X., X.C., V.U., B.G., A.N., and S.C. were involved in study design and interpretation of the analysis. A.L.F., K.J., R.S., J.L., A.C., W.X., V.U., B.G., A.N., and D.C.M. conducted data collection. A.L.F., J.L., A.C., D.K.E., W.X., X.C., V.U., B.G., A.N., D.C.M., and S.C. contributed to data analysis and interpretation. All authors contributed to the development of the initial draft of the manuscript, critically revised the manuscript, and approved the final version.

## DATA AVAILABILITY

Access to patient-level data presented in this article and supporting clinical documents with external researchers who provide methodologically sound scientific proposals will be available upon reasonable request for products or indications that have been approved by regulators in the relevant markets and subject to review from 24 months after study completion. Such requests can be made to Moderna Inc., 325 Binney Street, Cambridge, MA 02142, USA. A materials transfer and/or data access agreement with the sponsor will be required for accessing shared data. All other relevant data are presented in the paper. The protocols are available online at ClinicalTrials.gov: NCT06585241; NCT07089706.

## REFERENCES

1. Uraki R, Korber B, Diamond MS, Kawaoka Y. SARS-CoV-2 variants: biology, pathogenicity, immunity and control. Nature Reviews Microbiology 2025.

2. World Health Organisation. WHO TAG-VE Risk Evaluation for SARS-CoV-2 Variant Under Monitoring: LP.8.1. Accessed 11 February 2026.

3. Chen L, Kaku Y, Okumura K, et al. Virological characteristics of the SARS-CoV-2 LP.8.1 variant. The Lancet Infectious Diseases 2025; 25:e193.

4. Centers for Disease Control. COVID. Variants and Genomic Surveillance. Available at: https://www.cdc.gov/covid/php/variants/variants-and-genomic-surveillance.html. Accessed December 11 2022.

5. US Food & Drug Administration. COVID-19 Vaccines (2025-2026 Formula) for Use in the United States Beginning in Fall 2025. Available at: https://www.fda.gov/vaccines-blood-biologics/industry-biologics/covid-19-vaccines-2025-2026-formula-use-united-states-beginning-fall-2025. Accessed December 5 2025.

6. GISAID. hCoV-19 (SARS-CoV-2) Variants and Lineages. Available at: https://gisaid.org/hcov-19-variants-dashboard/.

7. CDC. Variants and Genomic Surveillance. Available at: https://www.cdc.gov/covid/php/variants/variants-and-genomic-surveillance.html. Accessed January 20 2026.

8. Jule Z, Römer C, Hossen T, et al. Evolution and Viral Properties of the SARS-CoV-2 BA.3.2 Subvariant. medRxiv 2025:2025.10.28.25338622.

9. Jackson LA, Anderson EJ, Rouphael NG, et al. An mRNA vaccine against SARS-CoV-2—preliminary report. N Engl J Med 2020; 383:1920–31.

10. Granados-Riveron JT, Aquino-Jarquin G. Engineering of the current nucleoside-modified mRNA-LNP vaccines against SARS-CoV-2. Biomedicine & Pharmacotherapy 2021; 142:111953.

11. Stewart-Jones GBE, Elbashir SM, Wu K, et al. Domain-based mRNA vaccines encoding spike protein N-terminal and receptor binding domains confer protection against SARS-CoV-2. Science Translational Medicine 2023; 15:eadf4100.

12. Chalkias S, Pragalos A, Akinsola A, et al. Safety and immunogenicity of SARS-CoV-2 spike receptor-binding domain and N-terminal domain mRNA vaccine. J Infect Dis 2025; 231:e754–e63.

13. Chalkias S, Dennis P, Petersen D, et al. Efficacy, immunogenicity, and safety of a next-generation mRNA-1283 COVID-19 vaccine compared with the mRNA-1273 vaccine (NextCOVE): results from a phase 3, randomised, observer-blind, active-controlled trial. Lancet Infect Dis 2025.

14. Moderna TX, Inc. MNEXSPIKE (COVID-19 vaccine, mRNA). Package insert. Vol. 2025: US Food and Drug Administration, 2025.

15. Nasir A, Lee D, Avena LE, et al. Predictive Modeling of Immune Escape and Antigenic Grouping of SARS-CoV-2 Variants. bioRxiv 2025:2025.05.28.656328.

16. Bloom J. SARS2-RBD-escape-calc,. Available at: https://raw.githubusercontent.com/jbloomlab/SARS2-RBDescape-calc/main/results/escape.csv. Accessed January 20 2026.

17. Yisimayi A, Song W, Wang J, et al. Repeated Omicron exposures override ancestral SARS-CoV-2 immune imprinting. Nature 2024; 625:148–56.

18. Cao Y, Jian F, Wang J, et al. Imprinted SARS-CoV-2 humoral immunity induces convergent Omicron RBD evolution. Nature 2023; 614:521–9.

19. Greaney AJ, Starr TN, Bloom JD. An antibody-escape estimator for mutations to the SARS-CoV-2 receptor-binding domain. Virus Evol 2022; 8:veac021.

20. WHO. Statement on the antigen composition of COVID-19 vaccines. Available at: https://www.who.int/news/item/18-12-2025-statement-on-the-antigen-composition-of-covid-19-vaccines. Accessed January 20 2026.

