## Supplement for "Immunogenicity and safety of LP.8.1 variant-containing mRNA COVID-19 vaccines"

### SUPPLEMENTARY METHODS

#### Inclusion and Exclusion Criteria for the mRNA-1273.251 or mRNA-1283.251 Study

Each participant was required to have met the following criteria for enrollment in the study:

- Individuals aged ≥65 years at the time of signing the informed consent or aged ≥12 to <65 years at the time of signing the informed consent with at least 1 risk factor for severe outcomes from COVID-19 (adapted from CDC 2025)
- Male or female assigned at birth, inclusive of all gender identities
- Investigator’s assessment that the participant understands and was willing and physically able to comply with protocol-mandated follow-up, including all procedures
- Contraceptive use by participants or the participants partners should be consistent with local regulations regarding the methods of contraception for those that participate in clinical trials
- Participants who were assigned female at birth or could become pregnant:
    - Had a negative pregnancy test at the screening visit and on the day of vaccination prior the administration of the vaccine dose on Day 1
    - Had practiced adequate contraception or had abstained from all activities that could result in pregnancy for at least 28 days prior to the first dose (Day 1). Adequate female contraception was defined as consistent and correct use of a local health authority approved contraceptive method in accordance with the product label

    - Had agreed to continue adequate contraception through 28 days following vaccine administration
- Capable of giving signed informed consent which includes compliance with the requirements and restrictions listed in the informed consent form and in the protocol

Participants who met any of the following criteria were excluded from the study:

- History of SARS-CoV-2 infection within 6 months prior to enrollment
- Acutely ill or febrile (temperature ≥38.0°C [≥100.4°F]) prior to or at screening visit or Day 1. Participants meeting this criterion may be rescheduled within the screening window and will retain their initially assigned participant number
- History of a diagnosis or condition that, in the judgment of the Investigator, was clinically unstable or may affect participant safety, assessment of study endpoints, assessment of immune response, or adherence to study procedures. Clinically unstable is defined as a diagnosis or condition requiring changes in management or medication within the 60 days prior to screening and includes ongoing workup of an undiagnosed illness that could lead to a new diagnosis or condition
- Reported history of congenital or acquired immunodeficiency, immunosuppressive condition, asplenia, or recurrent severe infection. Certain immune-mediated conditions that are stable and well-controlled (e.g., Hashimoto thyroiditis) as well as those that do not require systemic immunosuppressants per exclusion criterion bullet 7, sub bullet 3 (e.g.,

asthma, psoriasis, or vitiligo) may be permitted at the discretion of the Investigator

- History of coagulopathy or bleeding disorder that is considered a contraindication to intramuscular (IM) injection or phlebotomy
- Any medical, psychiatric, or occupational condition, including reported history of drug or alcohol abuse, that, in the opinion of the Investigator, might pose additional risk due to participation in the study or could interfere with the interpretation of study results
- Receipt of the following:
    - COVID-19 vaccine within 6 months prior to enrollment
    - Any licensed non-COVID-19 vaccine within 28 days before or planned receipt within 28 days after the study intervention, except an influenza vaccine, which may be given 14 days before or after receipt of the study intervention
    - Systemic immunosuppressants for >14 days in total, within 180 days prior to the screening visit (for corticosteroids ≥10 mg/day of prednisone equivalent) or was anticipating the need for systemic immunosuppressive treatment at any time during participation in the study
    - Participants who received epidural or major joint intra-articular injections (e.g., knee, hip, shoulder) within 28 days prior to Day 1 or planned to receive them within 28 days after Day 1 were excluded. Inhaled, nasal, and topical steroids were allowed

    - Systemic immunoglobulins, long-acting biological therapies that affect immune responses (e.g., infliximab) or blood products within 90 days prior to the screening visit or planned to receive them during the study
- History of anaphylaxis or severe hypersensitivity reaction requiring medical intervention after receipt of any mRNA vaccine or therapeutic or any components of an mRNA vaccine or therapeutic
- History of myocarditis, pericarditis, or myopericarditis within 90 days prior to the screening visit. Participants who had not returned to baseline after their convalescent period were also excluded
- History of Guillain-Barrè syndrome
- Had donated ≥450 mL of blood products within 28 days prior to the screening visit or planned to donate blood products within 28 days after the study injection
- Had participated in an interventional clinical study within 28 days prior to the screening visit based on the medical history interview or planned to do so while participating in this study. Interventions such as counseling, biofeedback, and cognitive therapy were not exclusionary
- Is working or has worked as study personnel or is an immediate family member or household member of study personnel, study site staff, or Sponsor personnel or is someone directly involved with the conduct of the study

**Pseudovirus-based neutralization assay**

**Immunogenicity Analyses**

Serum neutralizing antibodies (nAbs) against the vaccine-matched SARS-CoV-2 variant (LP.8.1; primary endpoint) and nAbs against SARS-CoV-2 variants of concern or new variants (LP.8.1, BA.3.2.2, XFG, NB.1.8.1; ad hoc endpoint) following vaccination were measured by the pseudovirus-based neutralization assay. Lentiviral particles displaying the SARS-CoV-2 Spike protein of interest (LP.8.1, BA.3.2.2, XFG, NB.1.8.1) and encoding firefly luciferase were used to infect 293T/ACE2 cells in the presence of serially diluted sera/antibodies. Infection was quantified by relative luminescence units (RLU). Neutralization titers were reported as $ID_{50}$ and $ID_{80}$, defined as the serum dilutions reducing background-corrected RLU by 50% or 80%, respectively, relative to virus-only controls (cell-only wells subtracted). Seroresponse was defined as a change from below the LLOQ to ≥4× LLOQ, or at least a 4-fold rise if the baseline was ≥ the LLOQ.

### SUPPLEMENTARY TABLES

**Table S1. Neutralizing antibody titers against LP.8.1 by age group at baseline and Day 29, per protocol primary analysis**

| | mRNA-1273.251<br>N = 52 | mRNA-1283.251<br>N = 98 |
|---|---|---|
| **Age group ≥12 to <65 years** | | |
| Baseline, n[a] | 52 | 98 |
| GMT (95% CI)[b] | 87.6 (55.1-139.2) | 62.0 (43.3-88.7) |
| Day 29, n[a] | 52 | 97 |
| GMT (95% CI)[b] | 2300.9 (1443.4-3667.8) | 3230.3 (2246.4-4645.1) |
| GMFR (95% CI)[b] | 26.3 (15.6-44.3) | 53.0 (36.7-76.4) |
| SRR, n/N1 (%)[c,d] | 42/52 (86.5) | 86/97 (88.7) |
| **Age group ≥65 years** | | |
| Baseline, n[a] | 49 | 60 |
| GMT (95% CI)[b] | 90.4 (58.1-140.5) | 91.3 (59.5-140.1) |
| Day 29, n[a] | 49 | 58 |
| GMT (95% CI)[b] | 1394.6 (895.9-2170.9) | 3433.9 (2194.0-5374.5) |
| GMFR (95% CI)[b] | 15.4 (9.5-25.0) | 36.7 (23.1-58.4) |
| SRR, n/N1 (%)[c,d] | 41/49 (83.7) | 52/58 (89.7) |

GMT, geometric mean titer; GMFR, geometric mean fold-rise; LLOQ, lower limit of quantification; nAb, neutralizing antibody; SRR, seroresponse rate; ULOQ, upper limit of quantification.

Data cutoff date for immunogenicity: October 23, 2025 (mRNA-1273.251); October 31, 2025 (mRNA-1283.251).

Antibody values reported as <LLOQ are replaced by 0.5 x LLOQ. Values >ULOQ are replaced by the ULOQ. mRNA-1273.251 nAb LLOQ 35, ULOQ 38606. mRNA-1283.251 nAb LLOQ 35, ULOQ 38606.

[a]n represents the number of participants with non-missing data at baseline (Day 1) or post-baseline (Day 29).

[b]95% CI is calculated based on the t-distribution of the log-transformed values or the difference in the log-transformed values for GMT and GMFR, respectively, then back transformed to the original scale for presentation.

[c]Seroresponse at a participant level is defined as a change from <LLOQ to equal or above 4 x LLOQ, or at least a 4-fold rise if baseline is equal to or >LLOQ.

[d]Percentages are based on N1 (number of participants with non-missing data at baseline and Day 29).

### SUPPLEMENTARY FIGURES

**Figure S1. Neutralizing antibody responses elicited by (A) mRNA-1273.251 and (B) mRNA-1283.251 against vaccine-matched (LP.8.1) and emerging variants (BA.3.2.2, XFG, and NB.1.8.1) stratified by age**

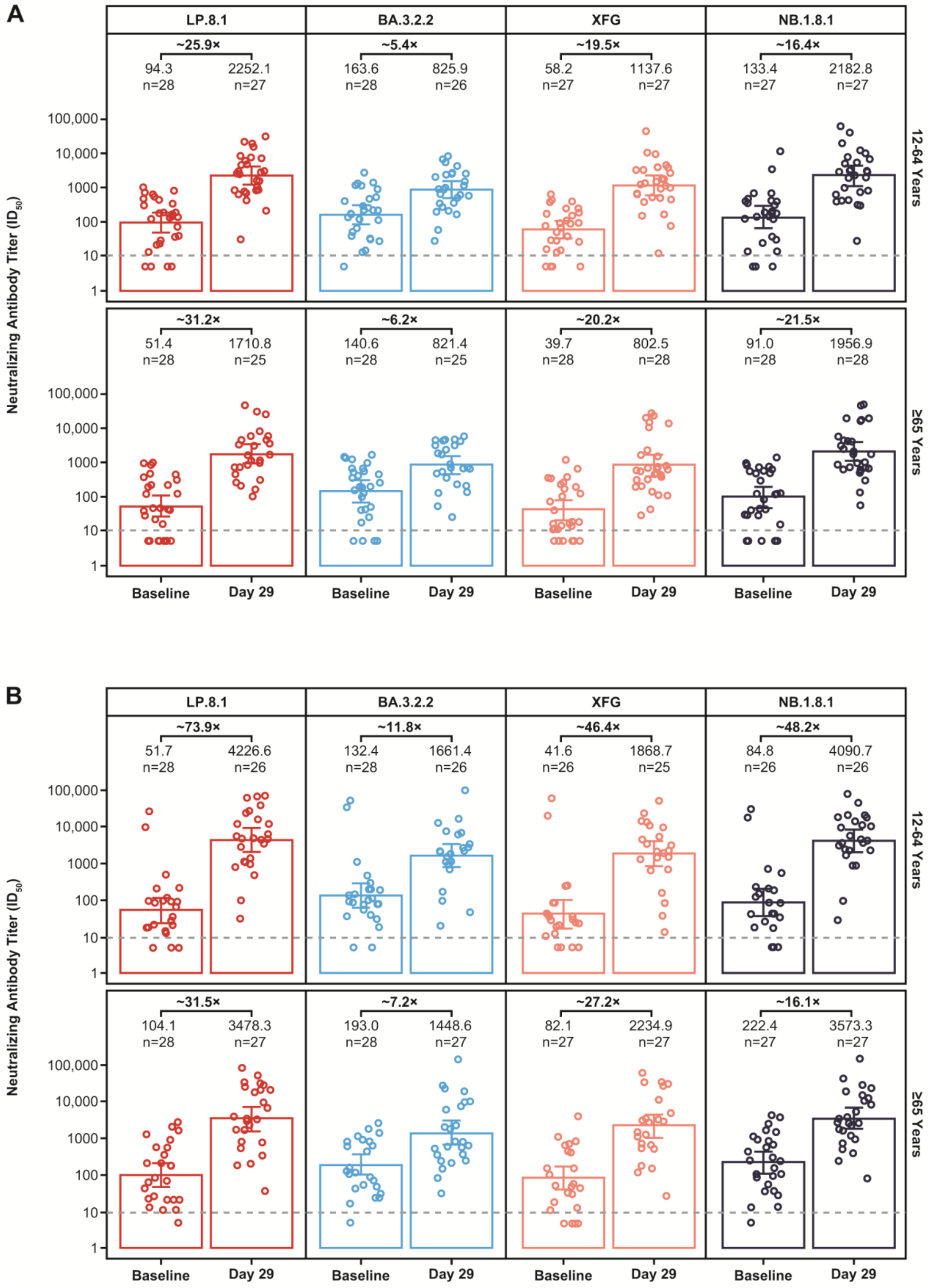


nAb, neutralizing antibody; $ID_{50}$, 50% inhibitory dose.

Dashed lines represent limit of detection.
